# Increased reproductive tract infections among secondary school girls during the COVID-19 pandemic: associations with pandemic related stress, mental health, and domestic safety

**DOI:** 10.1101/2024.02.17.24302973

**Authors:** Supriya D. Mehta, Debarghya Nandi, Fredrick Otieno, Garazi Zulaika, Elizabeth Nyothach, Walter Agingu, Runa Bhaumik, Linda Mason, Anna Maria van Eijk, Penelope A Phillips-Howard

## Abstract

**Background:** Kenya, like many countries, shuttered schools during COVID-19, with subsequent increases in poor mental health, sexual activity, and pregnancy. We sought to understand how the COVID-19 pandemic may mediate risk of reproductive tract infections. We hypothesized that greater COVID-19 related stress would mediate risk via mental health, feeling safe inside the home, and sexual exposure, given the pandemic mitigation-related impacts of school closures on these factors.

**Methods:** We analyzed data from a cohort of 436 girls enrolled in secondary school in rural western Kenya. Baseline, 6-, 12-, and 18-month study visits occurred April 2018 – December 2019 (pre-COVID), and 30-, 36-, and 48-month study visits occurred September 2020 – July 2022 (COVID period). At study visits, participants self-completed a survey for sociodemographics and sexual practices, and provided self-collected vaginal swabs for Bacterial vaginosis (BV) testing, with STI testing at annual visits. COVID-related stress was measured with a standardized scale and dichotomized at highest quartile. Mixed effects modeling quantified how BV and STI changed over time, and longitudinal mediation analysis quantified how the relationship between COVID-19 stress and increased BV was mediated.

**Findings:** BV and STI prevalence increased from 12.1% and 10.7% pre-COVID to 24.5% and 18.1% during COVID, respectively. This equated to a 26% (95% CI 1.00 – 1.59) and 36% (95% CI 0.98 – 1.88) increased relative prevalence of BV and STIs, respectively, in the COVID-19 period compared to pre-COVID, adjusted for numerous sociodemographic and behavioral factors. Higher COVID-related stress was associated with elevated depressive symptoms and feeling less safe inside the home, which were each associated with increased likelihood of having a boyfriend. In longitudinal mediation analyses, the direct effect of COVID-related stress on BV was small and non-significant, indicating increased BV was due to the constellation of factors that were impacted during the COVID-pandemic.

**Conclusions:** In this cohort of adolescent girls, BV and STIs increased following COVID-related school closures. These results highlight modifiable factors to help maintain sexual and reproductive health resiliency, such as anticipating and mitigating mental health impacts, domestic safety concerns, and maintaining sexual health services to prevent and treat reproductive tract infections.

## 1. INTRODUCTION

The World Health Organization (WHO) declared the end of the COVID-19 pandemic as a public health emergency on May 5, 2023. To help stem the initial impact of COVID-19, Kenya, like many countries around the world, shuttered schools on March 16, 2020, and put in place restrictions and curfews related to travel and social distancing [1]. Schools were partially reopened on October 19, 2020, and fully reopened on January 4, 2021. The closure of schools, curfews, and restrictions on travel and public gatherings were costly to the economy and disruptive to the social fabric of Kenyans. The World Bank reported an estimated 2 million Kenyans were newly forced into poverty due to the pandemic [2]. The tangible impacts of the economic decline led to increases in job loss and food insecurity, especially during the first 6 months of the pandemic, and this fall-out extended to many low- and middle-income countries [3].

During the school closures, there were increases in adolescent pregnancy observed in our cohort of secondary schoolgirls in western Kenya [4]. Among these secondary schoolgirls, the pandemic period was associated with a 3-fold higher risk of school dropout relative to pre-COVID-19 learners. This was a global phenomenon; school closures due to COVID-19 led to permanent school dropout stemming from exacerbated and expanded poverty, added responsibilities of employment, domestic work, and childcare, and pregnancy and marriage [5]. In meta-analyses of worldwide data, there were increases in depressive symptoms and anxiety among children and adolescents, with greater prevalence and severity seen among girls and older children [6,7]. Evidence also emerged showing increases in sexual offenses against children coincident with lockdowns, curfews and school closures [8].

Information is emerging on the impact of the COVID-19 pandemic on rates of sexually transmitted infections (STIs), which disproportionately affect adolescent girls and young women. While there were some decreases in STIs during the initial lockdown phases likely due to underreporting and limited access to services [9,10], STIs rebounded at increasing rates over the pre-COVID period in the United States [11,12], China [13], and Spain [14]. Maintaining reproductive tract health is a public health and clinical priority. Bacterial vaginosis (BV) increases the risk of HIV acquisition 1.6-fold [15] and non-ulcerative STIs increase the risk of HIV acquisition 3- to 5-fold [16]. This is especially relevant in western Kenya, where the prevalence of HIV in the general adult population ranges from 16% (Kisumu County) to 21% (Siaya County) [17]. BV and STIs are also associated with increased risk of adverse pregnancy outcomes, such as preterm birth and premature rupture of membranes [18], which already occur at increased rates among adolescent and young women (AGYW) [19].

In this analysis, we describe how the prevalence of reproductive tract infections changed in a cohort of Kenyan secondary schoolgirls from before to during and after the COVID-19 pandemic. We also sought to understand how the COVID-19 pandemic could mediate subsequent risk of reproductive tract infections. We hypothesized that greater COVID-19 related stress would mediate risk via mental health, safety in the home, and sexual exposure, given the pandemic mitigation-related impacts of school closures on these factors. Quantifying the extent to which these factors contribute to reproductive tract infection risk can help with decision-making around intervention development and prioritization.

## 2. METHODS

This study was approved by the institutional review boards of the Kenya Medical Research Institutes Scientific Ethics Review Unit (KEMRI, SERU #3215), Maseno University Ethics Review Committee (MUERC, MSU/DRPI/MUERC/01021/21), (Liverpool School of Tropical Medicine (LSTM, #15-005), and University of Illinois at Chicago (UIC, #2017-1301). Written informed consent was obtained for all participants, with written assent and guardian consent obtained for non-emancipated minors.

### 2.1 Study Design and Participants

Data for this analysis came from the Cups and Community Health (CaCHe) study [20,21], a sub-set of participants within the Cups or Cash for Girls (CCG) trial (ClinicalTrials.gov NCT03051789). The CCG trial has been described in detail [22]. Briefly, CCG was a cluster-randomized controlled trial in which secondary schools were randomized into 4 arms (1:1:1:1): (1) provision of menstrual cups with training on safe cup use and care; (2) conditional cash transfer (CCT) based on ≥80% school attendance in previous term; (3) menstrual cup and CCT; and (4) usual practice. For the CaCHe study, we enrolled approximately 20% of girls in the cup only and control arms of the CCG trial. The CaCHe study was powered to detect a 25% reduced relative prevalence of BV for the cup arm compared to the control arm, over 6 study visits. After enrollment, CaCHe participants were followed every 6 months through 30 months for the trial analysis [20] (Figure 1). The 24-month study visit scheduled to occur May 2020 was missed due to the COVID-19 pandemic. The 42-month visit was missed due to a gap in funding.

**Figure 1.**
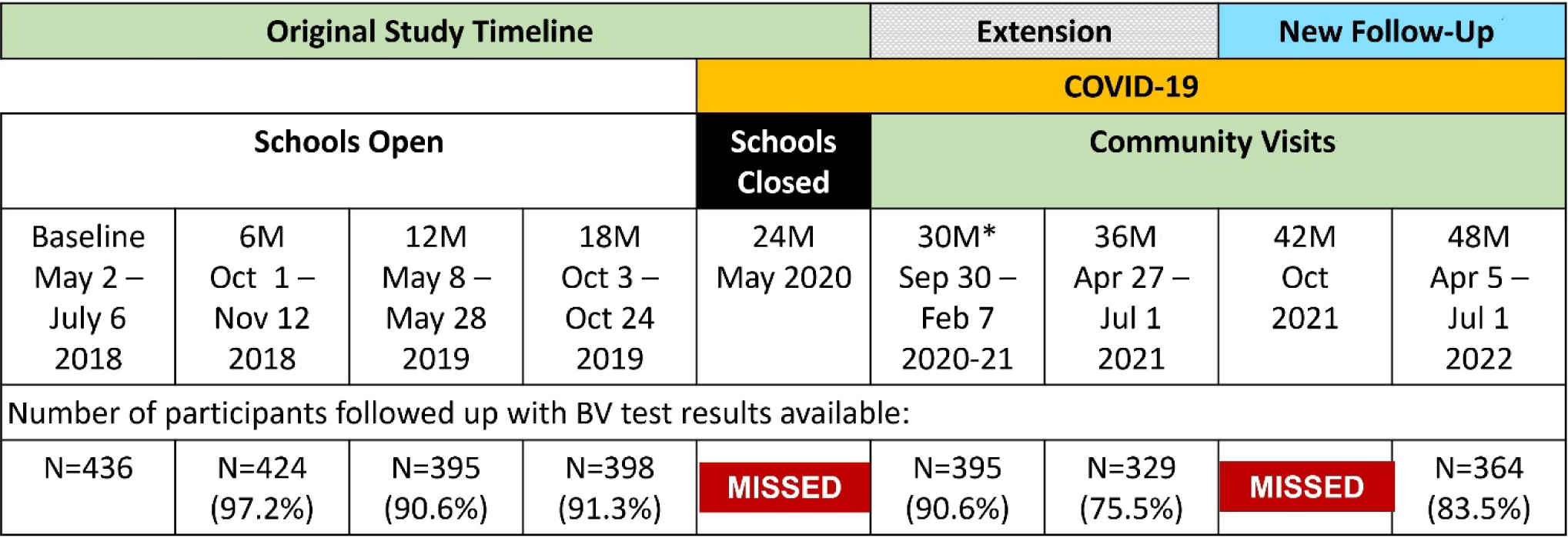
Timeline of CaCHe Study Visits. **Legend:** Baseline began May 2018, and study visits took place at schools every 6 months through October 2019 (18-month visit). The 24-month planned endline visit was cancelled due to COVID-19 restrictions and school closures. As schools were closed, we conducted the 30-month visit in the community, as were subsequent study visits due to participants completing school. With careful rebudgeting, we extended follow-up to 36-months, to continue capturing COVID-19 related impacts. At the 36-month visit, specimens were obtained from 329 participants; an additional 58 participants who had relocated were followed-up by telephone as the funding available did not support travel outside the area. Subsequent funding allowed for follow-up for all participants, including those who left the study area. *Due to COVID-19 precautions which limited the number of persons able to gather, the study period for the 30-month visit was extended, with 372 participants completed between September 30 – December 18, 2020, and an additional 25 participants between January 15 – February 7, 2021.

### 2.2 Data Collected

At each study visit, participants undertook a self-completed survey that collected information on sociodemographics, sexual practices, mental health, and menstrual practices. Participants were asked if they were sexually active in the past 6 months, and if they had been coerced or tricked to have sex in the past 6 months. Transactional sex was assessed through a series of questions that assessed sex in exchange for things (e.g., pads, money, school fees) or treatment (e.g., favors, employment). Girls were asked their marital status at each visit. Among those who reported being sexually active, they were asked whether it was with a boyfriend, husband, or partner. At the 18-month study visit, a separate question was added asking if participants were sexually active with someone who they consider to be a boyfriend, partner or husband in the past 6 months, and this variable was used for the 18-month visit onwards. Few participants reported being married until the 48-month visit (Table 1) and thus the variable used for analysis is referred to as having a “boyfriend” for simplicity. At the 12-, 30-, 36-, and 48-month visits, depressive symptoms were assessed using the 9-item Personal Health Questionnaire (PHQ-9); scores were dichotomized at 5 or higher for analyses, reflecting mildly elevated depressive symptoms [23]. The 12-month visit PHQ-9 score was applied at the baseline, 6-, and 18-month visits for longitudinal modeling due to lack of measure at these time points. Domestic safety was assessed at each time point during the COVID-19 period with a single question: “Since the curfews and school closures due to COVID-19, do you feel more safe or less safe inside your home?” with responses of “Less safe”, “The same”, “More safe”. For analysis, responses were dichotomized as “Less safe” vs. “The same or more safe”.

**Table 1.**
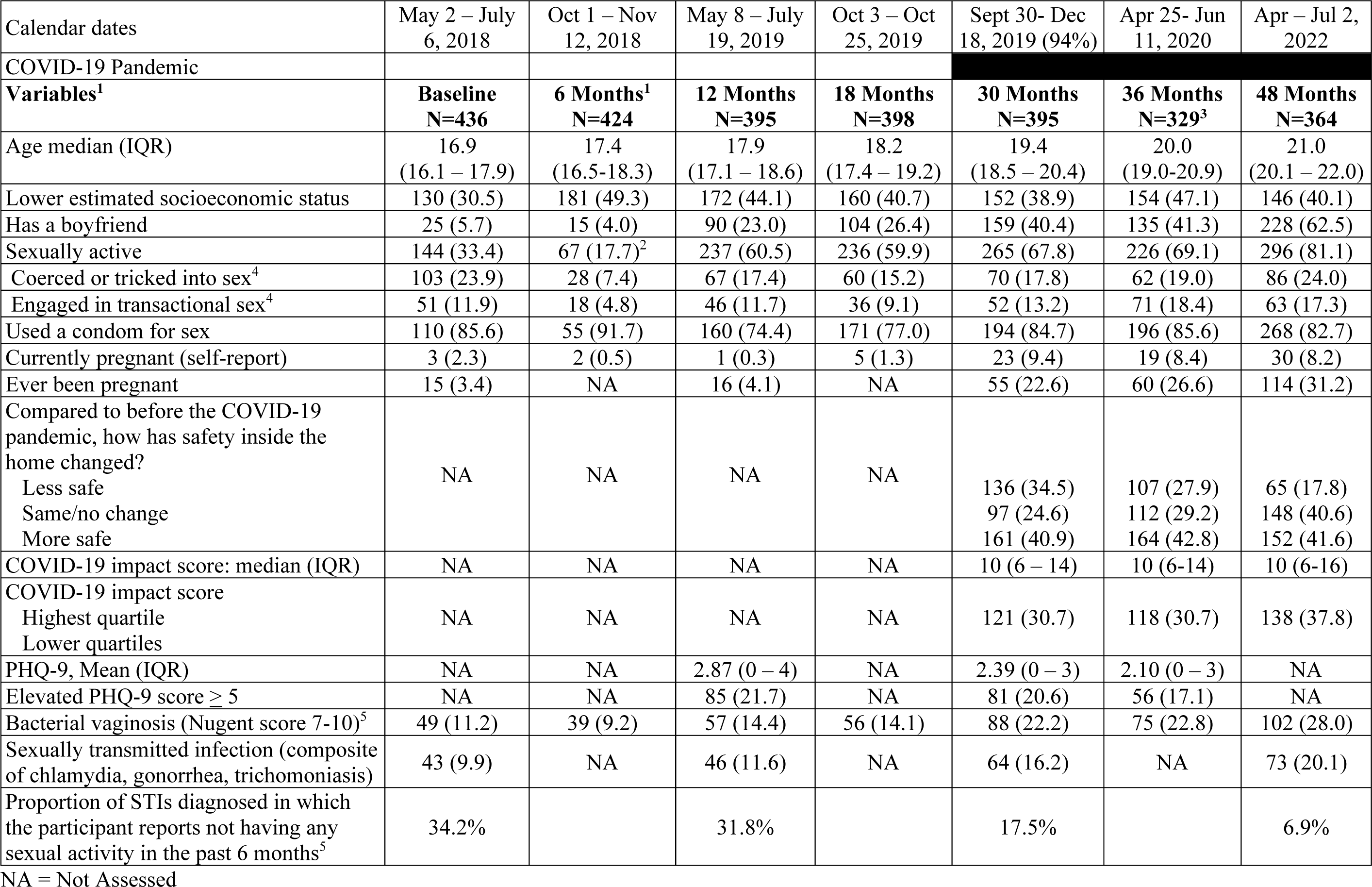
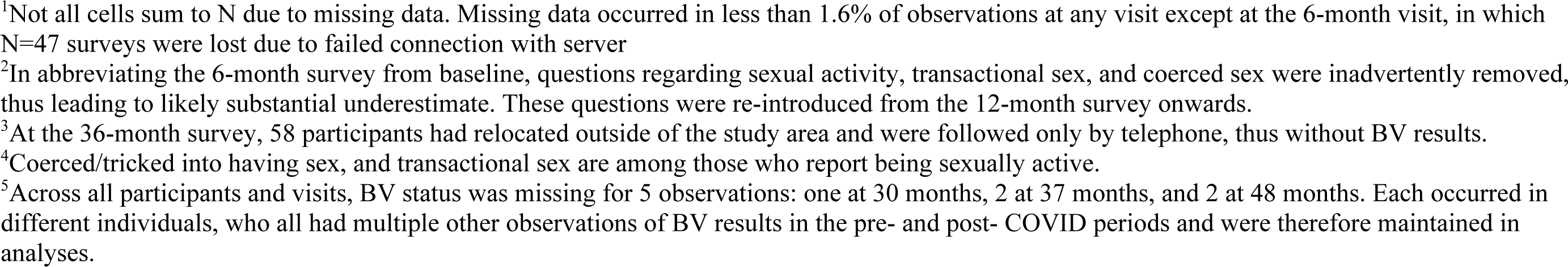
Distribution of Participant Characteristics by Study Time Point

| Calendar dates | May 2 – July 6, 2018 | Oct 1 – Nov 12, 2018 | May 8 – July 19, 2019 | Oct 3 – Oct 25, 2019 | Sept 30- Dec 18, 2019 (94%) | Apr 25- Jun 11, 2020 | Apr – Jul 2, 2022 |
| --- | --- | --- | --- | --- | --- | --- | --- |
| COVID-19 Pandemic |  |  |  |  |  |  |  |
| **Variables<sup>1</sup>** | **Baseline**<br>**N=436** | **6 Months<sup>1</sup>**<br>**N=424** | **12 Months**<br>**N=395** | **18 Months**<br>**N=398** | **30 Months**<br>**N=395** | **36 Months**<br>**N=329<sup>3</sup>** | **48 Months**<br>**N=364** |
| Age median (IQR) | 16.9<br>(16.1 – 17.9) | 17.4<br>(16.5-18.3) | 17.9<br>(17.1 – 18.6) | 18.2<br>(17.4 – 19.2) | 19.4<br>(18.5 – 20.4) | 20.0<br>(19.0-20.9) | 21.0<br>(20.1 – 22.0) |
| Lower estimated socioeconomic status | 130 (30.5) | 181 (49.3) | 172 (44.1) | 160 (40.7) | 152 (38.9) | 154 (47.1) | 146 (40.1) |
| Has a boyfriend | 25 (5.7) | 15 (4.0) | 90 (23.0) | 104 (26.4) | 159 (40.4) | 135 (41.3) | 228 (62.5) |
| Sexually active | 144 (33.4) | 67 (17.7)<sup>2</sup> | 237 (60.5) | 236 (59.9) | 265 (67.8) | 226 (69.1) | 296 (81.1) |
| Coerced or tricked into sex<sup>4</sup> | 103 (23.9) | 28 (7.4) | 67 (17.4) | 60 (15.2) | 70 (17.8) | 62 (19.0) | 86 (24.0) |
| Engaged in transactional sex<sup>4</sup> | 51 (11.9) | 18 (4.8) | 46 (11.7) | 36 (9.1) | 52 (13.2) | 71 (18.4) | 63 (17.3) |
| Used a condom for sex | 110 (85.6) | 55 (91.7) | 160 (74.4) | 171 (77.0) | 194 (84.7) | 196 (85.6) | 268 (82.7) |
| Currently pregnant (self-report) | 3 (2.3) | 2 (0.5) | 1 (0.3) | 5 (1.3) | 23 (9.4) | 19 (8.4) | 30 (8.2) |
| Ever been pregnant | 15 (3.4) | NA | 16 (4.1) | NA | 55 (22.6) | 60 (26.6) | 114 (31.2) |
| Compared to before the COVID-19 pandemic, how has safety inside the home changed? |  |  |  |  |  |  |  |
| Less safe | NA | NA | NA | NA | 136 (34.5) | 107 (27.9) | 65 (17.8) |
| Same/no change |  |  |  |  | 97 (24.6) | 112 (29.2) | 148 (40.6) |
| More safe |  |  |  |  | 161 (40.9) | 164 (42.8) | 152 (41.6) |
| COVID-19 impact score: median (IQR) | NA | NA | NA | NA | 10 (6 – 14) | 10 (6-14) | 10 (6-16) |
| COVID-19 impact score |  |  |  |  |  |  |  |
| Highest quartile | NA | NA | NA | NA | 121 (30.7) | 118 (30.7) | 138 (37.8) |
| Lower quartiles |  |  |  |  |  |  |  |
| PHQ-9, Mean (IQR) | NA | NA | 2.87 (0 – 4) | NA | 2.39 (0 – 3) | 2.10 (0 – 3) | NA |
| Elevated PHQ-9 score $\geq 5$ | NA | NA | 85 (21.7) | NA | 81 (20.6) | 56 (17.1) | NA |
| Bacterial vaginosis (Nugent score 7-10)<sup>5</sup> | 49 (11.2) | 39 (9.2) | 57 (14.4) | 56 (14.1) | 88 (22.2) | 75 (22.8) | 102 (28.0) |
| Sexually transmitted infection (composite of chlamydia, gonorrhea, trichomoniasis) | 43 (9.9) | NA | 46 (11.6) | NA | 64 (16.2) | NA | 73 (20.1) |
| Proportion of STIs diagnosed in which the participant reports not having any sexual activity in the past 6 months<sup>5</sup> | 34.2% |  | 31.8% |  | 17.5% |  | 6.9% |

NA = Not Assessed
<sup>1</sup>Not all cells sum to N due to missing data. Missing data occurred in less than 1.6% of observations at any visit except at the 6-month visit, in which N=47 surveys were lost due to failed connection with server <sup>2</sup>In abbreviating the 6-month survey from baseline, questions regarding sexual activity, transactional sex, and coerced sex were inadvertently removed, thus leading to likely substantial underestimate. These questions were re-introduced from the 12-month survey onwards. <sup>3</sup>At the 36-month survey, 58 participants had relocated outside of the study area and were followed only by telephone, thus without BV results. <sup>4</sup>Coerced/tricked into having sex, and transactional sex are among those who report being sexually active. <sup>5</sup>Across all participants and visits, BV status was missing for 5 observations: one at 30 months, 2 at 37 months, and 2 at 48 months. Each occurred in different individuals, who all had multiple other observations of BV results in the pre- and post- COVID periods and were therefore maintained in analyses.

Socioeconomic status at baseline was measured using questions related to household possessions, as reported in [24]. To reduce participant burden and for relevance to menstrual hygiene, only latrine type and water source used at home were asked at each subsequent 6 month visit and served as proxy for socioeconomic status (SES) as a time-updated measure. Latrine type was scaled 1 to 4 points (bush/field = 1, traditional pit = 2, improved pit = 3, flush toilet = 4) and water source was scaled 1 to 4 points (surface water = 1, borehole =2, rainwater = 3, piped water = 4) for a total of 8 points, with higher score reflecting greater improvement. This proxy SES score was dichotomized at 3 or less (lower) vs. 4 to 8 (higher). As compared to baseline SES using the more comprehensive assessment of household possessions, 79% of participants with higher proxy SES (4 to 8 score) also had higher SES by full assessment, and 42% of those with lower proxy SES (2-3 score) had lower SES by full assessment (tetrachoric rho = 0.54). School level water, sanitation, and hygiene (WASH) score at baseline was measured using previously described methods [23]; though schools were closed during the COVID-19 pandemic, this cluster-level variable was retained due to its relevance to area-level SES.

#### COVID-related stress

Stress related to the COVID-19 pandemic and mitigation measures was assessed at the 30-, 36-, and 48-month visits using 8 questions covering physical and emotional reaction domains related to COVID-19 specific psychological distress [25]. These questions ask about changes in physical (e.g., sleeping, concentrating) and emotional experiences (e.g., feelings of anxiousness, worry). Response categories were simplified from five categories to “Agree”, “Don’t know/Not Sure”, and “Disagree”. COVID-related stress score was calculated by assigning 2 points per question if the participant agreed, 1 point if participants responded “Don’t know/Not sure” response, and 0 if participants disagreed. The frequency distribution of responses to individual items over time are in Supplemental Table 1. Cronbach’s alpha was 0.87 (30-month), 0.86 (36-month), and 0.90 (48-month).

#### BV and STI Testing and Treatment

At baseline and each 6-month study visit, participants were asked to take self-collected vaginal swabs to test for BV and STIs, as detailed previously [20]. BV testing was done at each 6-month visit, and STI testing was conducted at baseline, 12-, 30-, and 48-month visits. Self-collected swabs for BV were immediately smeared on glass slides by the study staff and checked for sufficiency by a laboratory technician in the field. Slides were transported to the laboratory and gram stained, followed by Nugent’s scoring for detection of BV (Nugent score 7-10) [26]. Infection with *Neisseria gonorrhoeae* (NG) and *Chlamydia trachomatis* (CT) were assessed via nucleic acid amplification test (GeneXpert, Cepheid, Sunnydale, California, United States) and *Trichomonas vaginalis* (TV) by rapid immunochromatographic assay (OSOM TV antigen detection assay (Sekisui, Lexington, MA, US). All participants were provided results, and those who tested positive for BV or STIs were offered antibiotic treatment, regardless of symptoms.

### 2.3 Statistical Analysis

This analysis had two components: (1) mixed effects modeling to quantify how BV and STI changed from the pre- and post-COVID periods, and (2) longitudinal mediation analysis to explain how the relationship between COVID-19 stress and increased risk of BV was mediated. Due to minimal missing data, analyses were conducted as complete case.

#### 2.3.1. Change in BV and STIs from Before to During COVID-19

BV (Nugent score 7-10 vs. 0-6) is used as the primary outcome due to availability at all study visits (except the 24 months missed visit) and for high correlation with sexual activity [21]. We examined the change in BV over time using generalized linear mixed effects models with Poisson distribution, with random effects for participant and cluster (school) and robust variance estimate. We examined the prevalence ratio of BV during the COVID-19 period (visits 30-through 48-months) as compared to pre-COVID (baseline through 18-months), and multivariable models adjusted for intervention status and *a priori* confounders [20]: baseline STI status, age, socioeconomic status, PHQ-9, and reports of having a boyfriend and engaging in transactional sex. As a secondary analysis, STIs were modeled using mixed effects models as described above. Mixed effects models were conducted in Stata/SE v17 (College Station, TX).

#### 2.3.2 Longitudinal mediation model

We conducted longitudinal mediation analysis to test our hypotheses that greater COVID-related stress mediated elevated depressive symptoms and feeling less safe inside the home, which in turn mediated the proximal exposure, of having a boyfriend (Figure 2). The variable “sexually active” itself was not included in the model, since it is in the causal pathway for STIs, and often for BV. The outcome for analysis was BV, as STI was measured only at annual study visits. We controlled for assigned intervention status as a known factor associated with BV [20], socioeconomic status and age as level 1 confounders (Figure 2), and engaging in transactional sex as a level 2 confounder (Figure 2). The mediation approach differs from multivariable adjustment (approach 1, above), in that the independent variable (COVID-related stress) is proposed to *influence* the mediator variables (depressive symptoms, domestic safety, having a boyfriend), which in turn influences the dependent variable (BV). In this way, the mediator variables seek to clarify *how* the independent and dependent variables are related.

**Figure 2.**
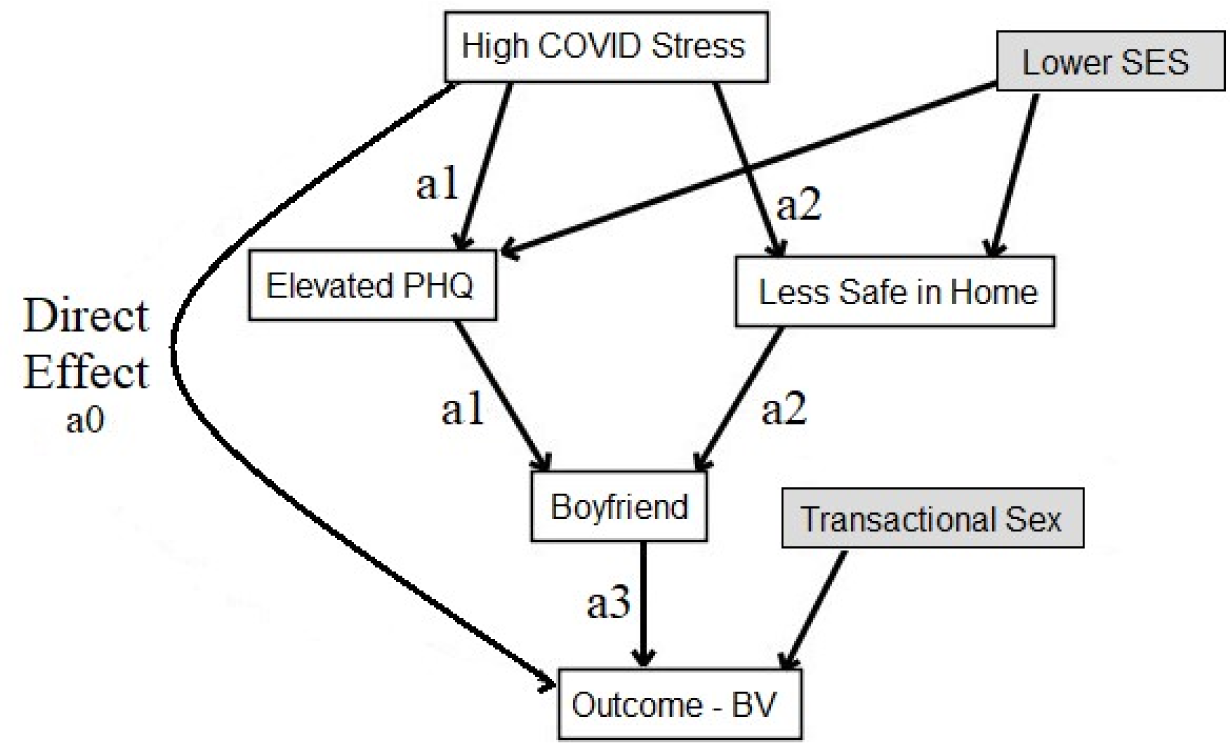
Conceptual model showing hypothesized linkages between COVID-related stress and Bacterial vaginosis. **Legend**: The hypothesized relationship between elevated COVID-related stress and BV (*direct effect, a0)* is depicted with pathways being mediated through level 1 mediators (*a1* elevated depressive symptoms (PHQ-9 ≥5) and *a2* feeling less safe inside the home compared to before the COVID pandemic) and level 2 mediator *a3* having a boyfriend. We hypothesized socioeconomic status confounded the association between elevated COVID-related stress and increased depressive symptoms and feeling less safe inside the home. We also hypothesized that transactional sex confounded the association between elevated COVID-related stress and BV. Assigned intervention status (menstrual cup arm or control arm) and participant age were also controlled for and are not depicted in the figure.

The longitudinal mediation analysis decomposed the overall impact of higher COVID-related stress on the risk of getting BV into multiple distinct pathways using a counterfactual framework for mediation [27,28]. We defined a generalized marginal structural model for nested counterfactuals where we directly parameterized each of the indirect effect pathways (Figure 2) through four distinct pathways: the *direct effect*, and indirect pathways *a1*, *a2*, and *a3*. The *a1* pathway encompasses all potential pathways involving elevated depressive symptoms (PHQ-9 ≥ 5), while *a2* encompasses those associated with feeling less safe at home compared to prior to the COVID-19 pandemic. Lastly, *a3* captures the influence of having a boyfriend on the outcome, BV. We derived parameter estimates of the direct and the indirect effects by regressing BV on COVID-19 related stress (the exposure) as well as all the other possible pathways, using a weighted approach and incorporating random intercepts [29]. Causal mediation analysis with a non-rare binary outcome often introduces some ambiguity due to the non-collapsibility of odds ratio, and we address this challenge using a log linear model with robust bootstrapping for standard error estimation [30]. We also report percentage of mediation effect. Longitudinal mediation was conducted in R (v4.1.13).

## 3. RESULTS

At baseline, participants were median age 16.9 years and were median age 21.0 years at the 48-month visit, in keeping with the 4-year span of time (Table 1). The proportion of participants reporting having a boyfriend and being sexually active increased over time, especially at the 12-, 30-, and 48-month visits. At the 30-month visit, when schools largely remained closed and curfews were still in place, over one-third (34.5%) of participants reported feeling less safe in their home as compared to before the COVID-19 pandemic and associated lockdowns; this decreased to 17.8% by the 48-month visit.

### 3.1. Distribution of Covariates by COVID-Related Stress

The median COVID-19 stress impact score was 10 at 30 months and remained similar across time points (Supplemental Table 1). There were higher rates of agreement with statements related to worry about getting infected with COVID, friend and family getting infected with COVID, or transmitting COVID to someone else, and lowest rates of agreement with difficulty sleeping, concentrating, and feeling overwhelmed.

### 3.2. Increase in BV and STIs in the COVID-19 period as compared to the pre-COVID period

Bacterial vaginosis increased from 11.2% at baseline to 14.1% at 18 months, and in the COVID-19 period, increased to 22.2% at 30 months and 28.1% by 48 months (Figure 3). Similarly, STI prevalence increased from 9.9% at baseline to 11.6% at 12 months, rising to 16.2% at the 30-month and to 20.1% by the 48-month visit (Figure 3). At each timepoint during the COVID-19 period, the prevalence of BV was greater for participants with higher reported COVID-related stress, those with lower socioeconomic indicators, reports of having a boyfriend, engaging in transactional sex, elevated PHQ-9 score, and feeling less safe inside the home (Table 2). In multivariable mixed effects modeling, there was a 26% increased relative prevalence of BV in the COVID-19 period compared to pre-COVID (adjusted prevalence ratio [aPR] = 1.26; 95% CI: 1.00 – 1.59; Table 3), adjusted for assigned intervention status; baseline school-level WASH score, socioeconomic status, sexual activity, and STI status; and time-varying age, socioeconomic status, elevated PHQ-9 score, engaging in transactional sex, and having a boyfriend. Similarly, there was a 36% increased prevalence ratio of STI in the COVID-19 period compared to the pre-COVID period (aPR = 1.36; 95% CI: 0.98 – 1.88) (Table 3).

**Figure 3.**
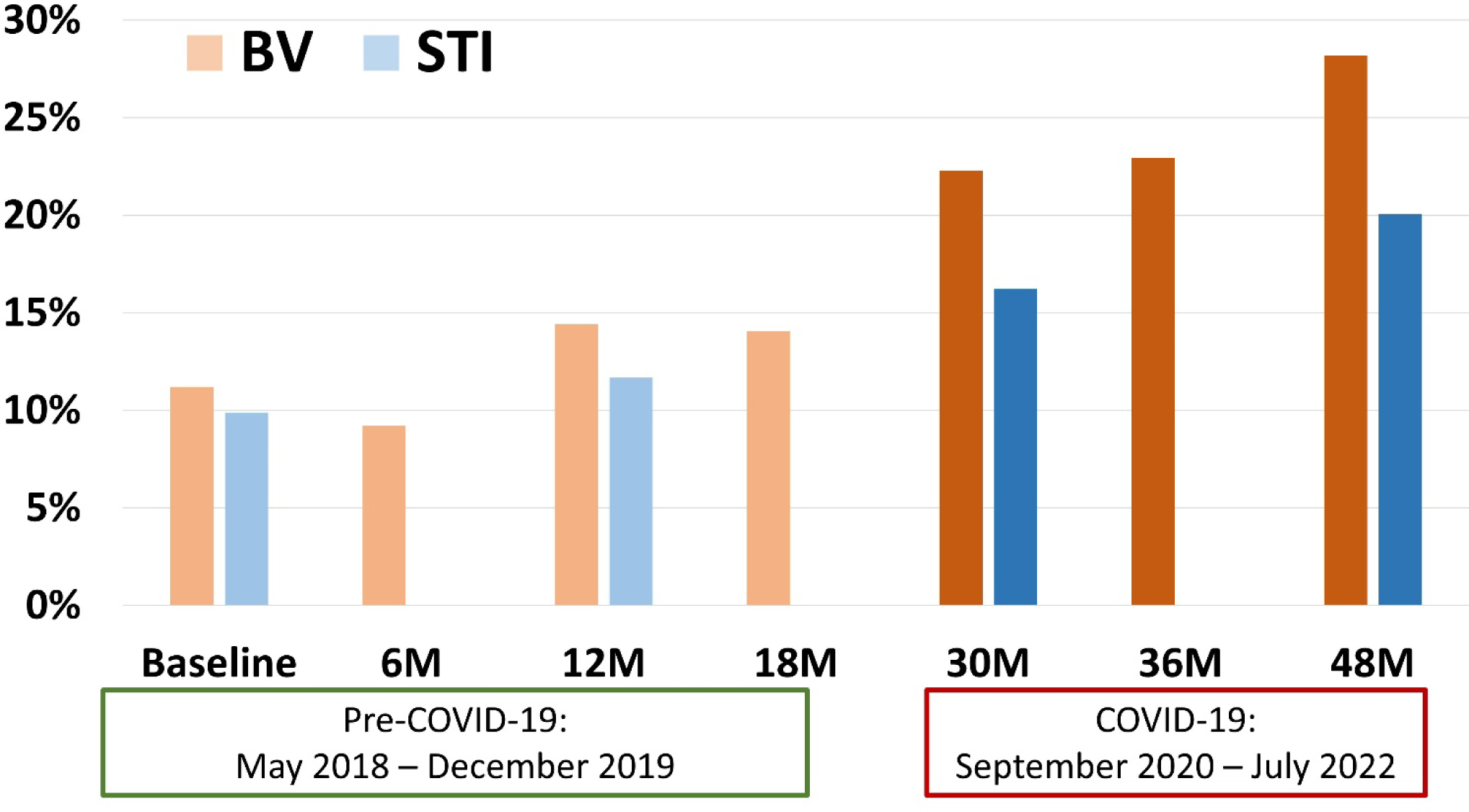
Prevalence of BV and STIs over time before and during COVID-19. Legend: The y-axis shows the prevalence of BV (orange) and STIs (blue) over study visits (x-axis). The pre-COVID and COVID-19 periods are indicated with the dates underneath the x-axis and darker intensity of coloring in the visits occurring during the COVID-19 period.

**Table 2.**
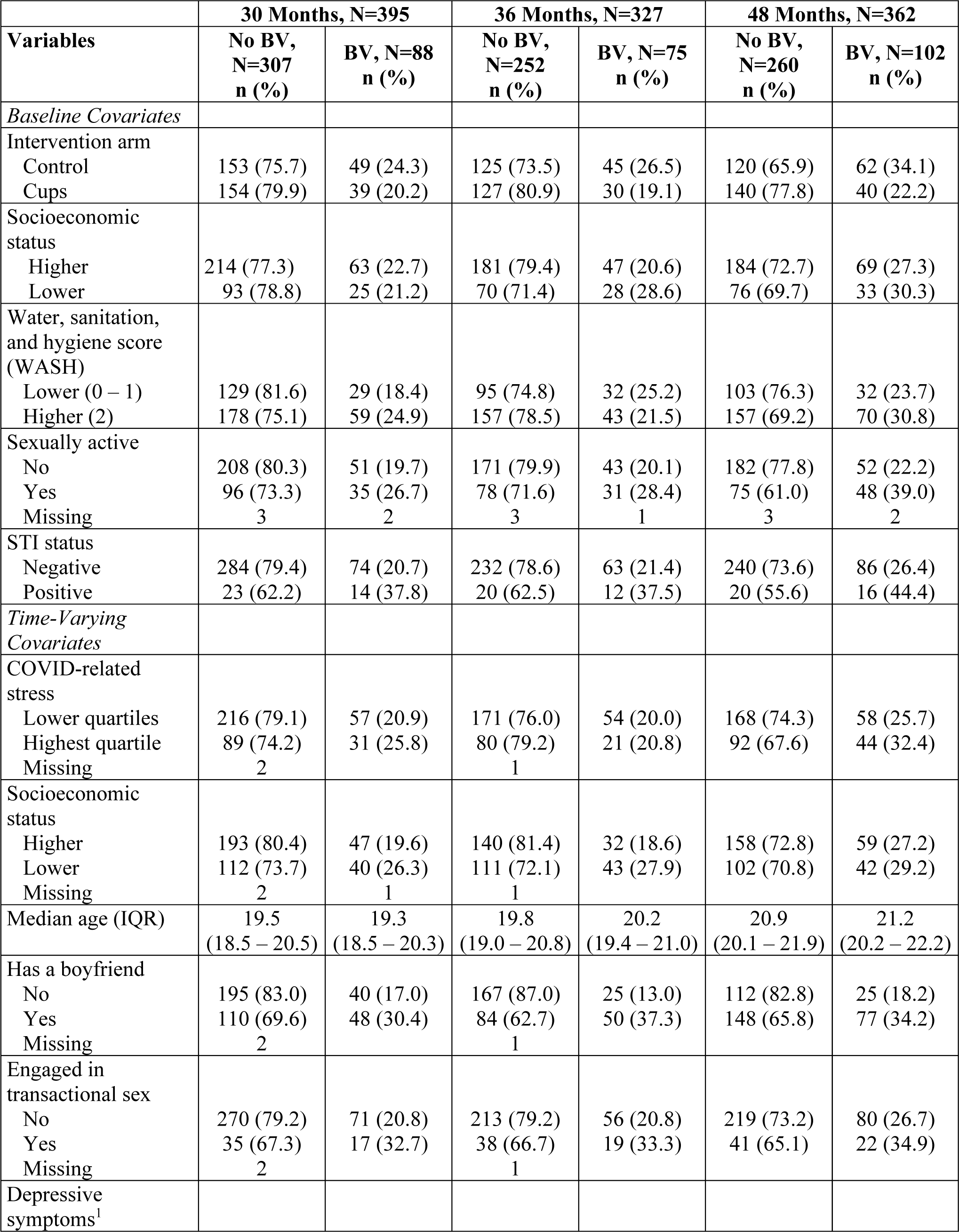
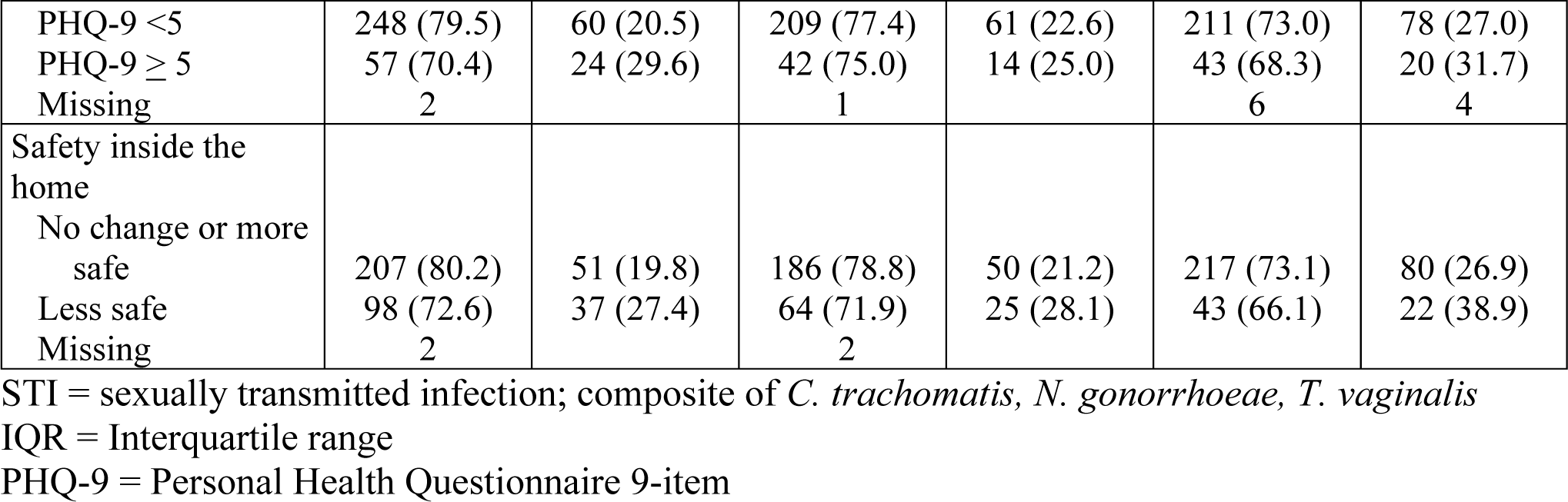
Distribution of COVID-19 related stress and hypothesized mediating factors by time point and Bacterial vaginosis (BV) status.

|  | **30 Months, N=395** |  | **36 Months, N=327** |  | **48 Months, N=362** |  |
| --- | --- | --- | --- | --- | --- | --- |
| **Variables** | **No BV,**<br>**N=307**<br>**n (%)** | **BV, N=88**<br>**n (%)** | **No BV,**<br>**N=252**<br>**n (%)** | **BV, N=75**<br>**n (%)** | **No BV,**<br>**N=260**<br>**n (%)** | **BV, N=102**<br>**n (%)** |
| *Baseline Covariates* |  |  |  |  |  |  |
| Intervention arm |  |  |  |  |  |  |
| Control | 153 (75.7) | 49 (24.3) | 125 (73.5) | 45 (26.5) | 120 (65.9) | 62 (34.1) |
| Cups | 154 (79.9) | 39 (20.2) | 127 (80.9) | 30 (19.1) | 140 (77.8) | 40 (22.2) |
| Socioeconomic status |  |  |  |  |  |  |
| Higher | 214 (77.3) | 63 (22.7) | 181 (79.4) | 47 (20.6) | 184 (72.7) | 69 (27.3) |
| Lower | 93 (78.8) | 25 (21.2) | 70 (71.4) | 28 (28.6) | 76 (69.7) | 33 (30.3) |
| Water, sanitation, and hygiene score (WASH) |  |  |  |  |  |  |
| Lower (0 – 1) | 129 (81.6) | 29 (18.4) | 95 (74.8) | 32 (25.2) | 103 (76.3) | 32 (23.7) |
| Higher (2) | 178 (75.1) | 59 (24.9) | 157 (78.5) | 43 (21.5) | 157 (69.2) | 70 (30.8) |
| Sexually active |  |  |  |  |  |  |
| No | 208 (80.3) | 51 (19.7) | 171 (79.9) | 43 (20.1) | 182 (77.8) | 52 (22.2) |
| Yes | 96 (73.3) | 35 (26.7) | 78 (71.6) | 31 (28.4) | 75 (61.0) | 48 (39.0) |
| Missing | 3 | 2 | 3 | 1 | 3 | 2 |
| STI status |  |  |  |  |  |  |
| Negative | 284 (79.4) | 74 (20.7) | 232 (78.6) | 63 (21.4) | 240 (73.6) | 86 (26.4) |
| Positive | 23 (62.2) | 14 (37.8) | 20 (62.5) | 12 (37.5) | 20 (55.6) | 16 (44.4) |
| *Time-Varying Covariates* |  |  |  |  |  |  |
| COVID-related stress |  |  |  |  |  |  |
| Lower quartiles | 216 (79.1) | 57 (20.9) | 171 (76.0) | 54 (20.0) | 168 (74.3) | 58 (25.7) |
| Highest quartile | 89 (74.2) | 31 (25.8) | 80 (79.2) | 21 (20.8) | 92 (67.6) | 44 (32.4) |
| Missing | 2 |  | 1 |  |  |  |
| Socioeconomic status |  |  |  |  |  |  |
| Higher | 193 (80.4) | 47 (19.6) | 140 (81.4) | 32 (18.6) | 158 (72.8) | 59 (27.2) |
| Lower | 112 (73.7) | 40 (26.3) | 111 (72.1) | 43 (27.9) | 102 (70.8) | 42 (29.2) |
| Missing | 2 | 1 | 1 |  |  |  |
| Median age (IQR) | 19.5<br>(18.5 – 20.5) | 19.3<br>(18.5 – 20.3) | 19.8<br>(19.0 – 20.8) | 20.2<br>(19.4 – 21.0) | 20.9<br>(20.1 – 21.9) | 21.2<br>(20.2 – 22.2) |
| Has a boyfriend |  |  |  |  |  |  |
| No | 195 (83.0) | 40 (17.0) | 167 (87.0) | 25 (13.0) | 112 (82.8) | 25 (18.2) |
| Yes | 110 (69.6) | 48 (30.4) | 84 (62.7) | 50 (37.3) | 148 (65.8) | 77 (34.2) |
| Missing | 2 |  | 1 |  |  |  |
| Engaged in transactional sex |  |  |  |  |  |  |
| No | 270 (79.2) | 71 (20.8) | 213 (79.2) | 56 (20.8) | 219 (73.2) | 80 (26.7) |
| Yes | 35 (67.3) | 17 (32.7) | 38 (66.7) | 19 (33.3) | 41 (65.1) | 22 (34.9) |
| Missing | 2 |  | 1 |  |  |  |
| Depressive symptoms<sup>1</sup> |  |  |  |  |  |  |
| PHQ-9 <5 | 248 (79.5) | 60 (20.5) | 209 (77.4) | 61 (22.6) | 211 (73.0) | 78 (27.0) |
| PHQ-9 ≥ 5 | 57 (70.4) | 24 (29.6) | 42 (75.0) | 14 (25.0) | 43 (68.3) | 20 (31.7) |
| Missing | 2 |  | 1 |  | 6 | 4 |
| Safety inside the home |  |  |  |  |  |  |
| No change or more safe | 207 (80.2) | 51 (19.8) | 186 (78.8) | 50 (21.2) | 217 (73.1) | 80 (26.9) |
| Less safe | 98 (72.6) | 37 (27.4) | 64 (71.9) | 25 (28.1) | 43 (66.1) | 22 (38.9) |
| Missing | 2 |  | 2 |  |  |  |

STI = sexually transmitted infection; composite of *C. trachomatis*, *N. gonorrhoeae*, *T. vaginalis*
IQR = Interquartile range
PHQ-9 = Personal Health Questionnaire 9-item

**Table 3.** Results of crude and multivariable adjusted mixed effects modeling: Prevalence ratio of Bacterial vaginosis and Sexually transmitted infections during COVID-19 as compared to pre-COVID^1^.

| **Bacterial vaginosis** | **Crude**<br>**N= 2,738**<br>**PR (95% CI)** | **Adjusted<sup>2</sup>**<br>**N=2,544**<br>**PR (95% CI)** |
| --- | --- | --- |
| During COVID-19 period vs. Pre-COVID | 2.00 (1.62 – 2.48) | 1.26 (1.00 – 1.59) |
| **Sexually transmitted infections**<br>(composite of testing positive for any of:<br>*C. trachomatis*, *N. gonorrhoeae*, *T. vaginalis*) | **Crude**<br>**N= 1,587**<br>**PR (95% CI)** | **Adjusted<sup>3</sup>**<br>**N=1,511**<br>**PR (95% CI)** |
| During COVID-19 period vs. Pre-COVID | 1.68 (1.40 – 2.02) | 1.36 (0.98 – 1.88) |

PR = Prevalence Ratio; CI = Confidence Interval
<sup>1</sup>The Pre-COVID study visits of Baseline through 18- months took place May 2018 through October 2019. Study visits at 30- through 48- months took place during the COVID-19 period, September 2020 through July 2022.
<sup>2</sup>Simultaneously adjusted for: Assigned intervention status; baseline water, sanitation, and hygiene score, socioeconomic status, sexual activity, and STI status; and time-varying age, socioeconomic status, PHQ-9 score, engaging in transactional sex, and having a boyfriend.
<sup>3</sup>Simultaneously adjusted for: Assigned intervention status; baseline water, sanitation, and hygiene score, socioeconomic status, sexual activity; and time-varying age, socioeconomic status, elevated PHQ-9 score, engaging in transactional sex, and having a boyfriend.

### 3.3. Results of Mediation model

We hypothesized that stress related to COVID-19 could be mediating the increased prevalence of reproductive tract infection through depressive symptoms and domestic safety, both adversely affected by the pandemic and potentially related to increased likelihood of having a boyfriend and thus sexual exposure. To demonstrate the plausibility of the hypothesized relationships, the association between COVID-related stress and mediators was examined with mixed effects models. As shown in Table 4, feeling less safe inside the home compared to before the COVID-19 pandemic (aPR = 2.16; 95% CI: 1.74 – 2.68) was elevated among participants with highest quartile COVID-related stress score, adjusted for numerous baseline and time-varying covariates, though elevated depressive symptoms was not. Adjusted for the same numerous baseline and time-varying confounders, having a boyfriend (level 2 mediator) was statistically significantly increased with both level 1 mediators: elevated depressive symptoms (aPR = 1.20; 95% CI: 1.08 – 1.33) and feeling less safe inside the home compared to before COVID-19 (aPR = 1.21; 95% CI: 1.06 – 1.39), but was not associated with COVID-related stress. In longitudinal mediation analysis, AGYW who reported higher COVID-19 related stress had a non-significant 11% increased prevalence ratio of BV (Table 5). Among the various pathways through which COVID-19 related stress could mediate the risk of BV, feeling less safe inside emerged as an important factor, contributing to 57.3% of the total effect. This was followed by the influence of having a boyfriend at 25.2%, and elevated depressive symptoms at 15.3%.

**Table 4.** Results of crude and multivariable adjusted mixed effects modeling: Prevalence Ratio of Level 1 Mediators associated with elevated COVID-related stress.

| Level 1 Mediators | Crude PR (95% CI) | Adjusted for Longitudinal Mediation<sup>1</sup> PR (95% CI) | Fully Adjusted<sup>2</sup> PR (95% CI) |
| --- | --- | --- | --- |
| **Elevated Depressive Symptoms: PHQ-9 greater than or equal to 5** | N=1,143 | N=1,084 | N=1,069 |
| Highest quartile COVID-related stress vs. lower COVID-related stress | 1.24<br>(0.86 – 1.80) | 1.25<br>(0.84 – 1.85) | 1.11<br>(0.83 – 1.49) |
| **Feels less safe inside the home compared to before COVID-19** | N=1,142 | N=1,083 | N=1,069 |
| Highest quartile COVID-related stress vs. lower COVID-related stress | 2.28<br>(1.79 – 2.19) | 2.27<br>(1.81 – 2.85) | 2.16<br>(1.74 – 2.68) |
| Level 2 Mediator | Crude PR (95% CI) | Adjusted for Longitudinal Mediation<sup>3</sup> PR (95% CI) | Fully Adjusted<sup>4</sup> PR (95% CI) |
| **Has a boyfriend** | N=1,143 | N=1,083 | N=1,069 |
| Highest quartile COVID-related stress vs. lower COVID-related stress | 1.16<br>(1.01 – 1.35) | 1.02<br>(0.89 – 1.16) | 0.98<br>(0.85 – 1.12) |
| Elevated depressive symptoms (PHQ $\geq 5$ vs. $< 5$) | | 1.20<br>(1.01 – 1.41) | 1.20<br>(1.08 – 1.33) |
| Feels less safe inside the home during COVID-19 as compared to before |  | 1.22<br>(1.03 – 1.45) | 1.21<br>(1.06 – 1.39) |

All models, including crude models, are time-adjusted.
PR = Prevalence Ratio; CI = Confidence Interval
<sup>1</sup> Simultaneously adjusted for: Assigned intervention status and time, and level 1 confounders (time-varying socioeconomic status and age).
<sup>2</sup> Simultaneously adjusted for: Assigned intervention status; baseline water, sanitation, and hygiene score, socioeconomic status, sexual activity, and STI status; time and time-varying age, socioeconomic status, elevated PHQ-9 score (for safety inside the home model), feels less safe inside the home (for depressive symptoms model) engaging in transactional sex, and having a boyfriend.
<sup>3</sup> Simultaneously adjusted for: Assigned intervention status and time, level 1 confounders (time-varying socioeconomic status and age), level 1 mediators (depressive symptoms, feeling safe inside the home), and level 2 confounder (transactional sex).
<sup>4</sup> Simultaneously adjusted for: Assigned intervention status; baseline water, sanitation, and hygiene score, socioeconomic status, sexual activity, and STI status; time, and time-varying age, socioeconomic status, elevated PHQ-9 score, safe inside home, and engaging in transactional sex.

**Table 5.** Results of crude and variable adjusted longitudinal pathway model: Prevalence Ratio of Bacterial vaginosis (BV) for adolescent girls and young women with higher COVID-related stress versus less stress, decomposed into multiple pathways.

|  | **Crude**<br>**Model**<br>**PR**<br>**Estimate** | **Longitudinal Pathway**<br>**Model**<br>**PR**<br>**Estimate (95% CI)** | **Percentage of**<br>**Mediation** |
| --- | --- | --- | --- |
| **Direct Effect:** Elevated COVID-19 related stress and BV | 1.11 | 1.00 (0.91 – 1.09) | 2.2% |
| **Indirect Effect (a1)**<br>Association with BV mediated through elevated depressive symptoms (PHQ-9 ≥ 5) | | 1.02 (0.94 – 1.10) | 15.3% |
| **Indirect Effect (a2)**<br>Association with BV mediated through feeling less safe inside the home as compared to before COVID |  | 1.06 (0.98 – 1.14) | 57.3% |
| **Indirect Effect (a3)**<br>Association with BV mediated through having a boyfriend |  | 1.03 (0.95 – 1.11) | 25.2% |
| **Total Effect** | 1.11 | 1.11 | 100% |

PR = Prevalence Ratio; CI = Confidence Interval based on Bootstrapped Standard Error.

## 4. DISCUSSION

In this cohort of AGYW in western Kenya, the prevalence of BV and STIs increased over time, with greater increases observed during the period of COVID-19, immediately following school closures. The data captured in our cohort with time accruing before and during the COVID-19 pandemic enabled us to quantify this increase and evaluate reasons behind it. AGYW who were more greatly affected by the pandemic, as measured by COVID-related stress, were also more likely to report feeling less safe inside the home, and in turn, were more likely to have a boyfriend, which was proximal to having BV. Despite the association of these factors with increased COVID-related stress and with BV, and counter to our hypothesis, the direct effect of COVID-related stress on BV was small and non-significant, indicating the increased likelihood of BV was due to the constellation of factors that were impacted during the COVID-pandemic.

Having a boyfriend, the most proximal factor to BV and having greater likelihood of sexual exposure, may represent a coping mechanism in response to financial stress, feeling less safe in the home, or feelings of depression and anxiety. As reviewed in a study of sexual coping mechanisms during the COVID-19 pandemic, sexual connections and the intimacy entailed “can help cope with stressors and traumatic events” [31]. While not disaggregated by gender, the most frequently reported sexual strategies were sex as a source of pleasure and intimacy, to bond with and please a partner, to express care and strengthen the relationship, or to relieve stress and relax. A cross-sectional survey of Kenyan school going adolescents aged 13-19 living in Nairobi or the Coast region found high prevalences of anxiety (19.1%) and elevated depressive symptoms (19.1% with PHQ-9 ≥ 10), as well as high mean scores for emotional and behavioral problems [32]. Mbithi et al. also observed that living in unsafe neighborhoods, being physically forced to have sex, and drinking alcohol were associated with increased odds of depressive symptoms and anxiety. In a mobile phone-based survey of 2,224 adult-adolescent (age 10-19 years) pairs from Kisumu, Nairobi, and Coastal region, 36% of adolescents reported depressive symptoms, and this was associated with adult loss of income, which in turn was associated with household tensions and violence [33]. We observed marginal significance of elevated depressive symptoms among those with higher COVID-19 related stress, and there are other dimensions of mental health that we did not measure. As we continue follow-up of these AGYW, we have incorporated expanded measures of stress and anxiety. Our qualitative work will incorporate questions related to what extent and how sexual relationships may be serving as a coping response to stress, and how these AGYW respond to and cope with stress more broadly. This may provide insight for future interventions to provide AGYW with healthy coping strategies.

Alternatively, having a boyfriend may represent increased sexual exposure through financial need or coercion. Our qualitative study involving focus groups with study participants and men in their communities provide further insight [34]. AGYW expressed that there were increased drivers for sex during the COVID-19 lockdowns, school closures, and restrictions: increased poverty with concomitant pressures for transactional sex, including from men, peers, and parents; boredom and freedom to visit boyfriends; COVID-19 restrictions and curfews that left them having to spend nights with boyfriends; greater competition for boyfriends, leading to more risk taking (e.g., having sex).

Men expressed that providing material and financial support was helping adolescent girls and young women during the pandemic, and at the same time they had awareness that this enabled them a power differential in engaging girls to have sex or to have sex without condoms. In semi-structured in-depth interviews with 34 adolescent-caregiver dyads who had participated in the *Shamba Maisha* agricultural microfinance intervention in Kenya, AGYW receiving the intervention reported “no longer needing to engage in transactional sex or have multiple concurrent sexual partners as a way to meet their basic needs” [35]. Interviews revealed that girls felt less pressure for reciprocating for assistance with sex and that power differentials were reduced, by not having to rely on men for food and other needs. Although *Shamba Maisha* intervention took place prior to the pandemic, structural interventions addressing underlying drivers of sexual risk taking may sustainably reduce AGYW vulnerability to economic disruptions, such as those brought by the COVID-19 pandemic.

Feeling less safe in the home compared to before the COVID-19 pandemic was elevated among AGYW reporting higher levels of COVID-related stress and was associated with increased prevalence of BV. As noted, worldwide there were increases in all forms of violence against women and girls (including domestic violence, gender-based violence, and sexual assault), and this has been named the “Shadow Pandemic” by UN Women [36]. The conditions of the pandemic, especially during initial lockdowns and longer lasting restrictions on movement, led to financial stress, isolation with abusers, tighter living quarters, reduced access to health care, police and justice, and social services. UN Women called for national responses that provide psychosocial support to survivors, expanded services (e.g., shelters, hotlines, counseling), and “strong messages” that violence against women and girls will be met with legal response [37]. The COVID-19 pandemic demonstrated there is need and opportunity for virtual applications to provide services [38], though the applicability of virtual services remains challenging for rural and other areas with pre-existing limited infrastructural resources for technology and confidentiality, and limited or non-existent access to in-person services. While we did not measure whether feeling less safe was physical, emotional, or sexual, results such as ours reflecting the downstream impacts of domestic safety on reproductive health of AGYW highlights the need to invest in and ensure these services are functionally in place prior to or shortly following the next population level crisis. Interventions that are more likely to have significant and sustained impact on preventing gender-based violence are not quick or easy solutions, such as community-based and school-based promotion of gender equality, transforming gender stereotypes and discriminatory norms, reforming discriminatory laws, ensuring women’s access to formal wage employment and education, and increasing women’s access to financial security [39]. As Mehta and Seeley have previously written [40], the progress in sexual and reproductive health of the past 30 years is at critical juncture due to ongoing population level crises of pandemics, climate change, and conflict.

## Limitations

The longitudinal multilevel mediation analysis with multiple mediators at each level is quite complex, particularly when dealing with binary as opposed to continuous mediators and outcomes. With limited work in this area, defining an appropriate model with assumptions that satisfies all the data constraints is challenging. Marginal structural modelling is an effective method when it comes to modeling sequential mediators, but there is limited work that implements time-varying multi-level mediation. For this reason, we were selective in our choice of mediators. Subsequent analyses will examine whether BV and STI risks changed in the post-COVID time period. We believe our findings may generalize to AGYW who attended secondary school and live in similar setting, but the impacts of the COVID-19 pandemic on reproductive tract infections and drivers of this among those who did not attend school or live in different settings may be different.

## Conclusions

In this cohort of adolescent girls and young women, the prevalence of BV and STIs increased following school closures during the COVID-19 pandemic. Longitudinal analysis demonstrated that this increased risk was mediated by depressive symptoms and feeling less safe in the home, which led to increased likelihood of having boyfriends and thus sexual exposures. These results highlight specific modifiable factors that can be targeted by interventions during crises, to help maintain sexual and reproductive health resiliency in AGYW: mitigating mental health impacts and domestic safety concerns. Our results also indicate further research is needed to elucidate the benefits and risks of relying on boyfriends and sex partners to cope with these stressors, and how these relationships may be leveraged as a resource.

## Supporting information

Supplemental Table and STROBE

## Data Availability

All data produced in the present study are available upon reasonable request to the authors

