## Supplemental Table and STROBE for "Increased reproductive tract infections among secondary school girls during the COVID-19 pandemic: associations with pandemic related stress, mental health, and domestic safety"

**Supplemental Table 1. Distribution of responses to COVID-19 related stress questions[1] by study time point.**

| Please tell us whether you agree, disagree, or don't know for the following statements. | 30 Month visit, N=394 n (%) | 36 Month visit, N=327 n (%) | 48 Month visit, N=365 n (%) |
|---|---|---|---|
| **I am very worried about getting the coronavirus/COVID** | | | |
| Agree | 306 (77.7) | 259 (79.2) | 272 (74.5) |
| Disagree | 57 (14.5) | 55 (16.8) | 75 (20.6) |
| Don't know | 31 (7.9) | 13 (4.0) | 18 (4.9) |
| **I am very worried about my family or friends getting the coronavirus/COVID** | | | |
| Agree | 312 (79.2) | 267 (81.6) | 279 (76.4) |
| Disagree | 54 (13.7) | 45 (13.8) | 68 (18.3) |
| Don't know | 28 (7.1) | 15 (4.6) | 18 (4.9) |
| **I am very worried about giving someone else the coronavirus/COVID** | | | |
| Agree | 286 (72.6) | 234 (71.6) | 256 (70.1) |
| Disagree | 78 (19.8) | 74 (22.6) | 88 (24.1) |
| Don't know | 30 (7.6) | 19 (5.8) | 21 (5.8) |
| **I have had a hard time sleeping because of the coronavirus/COVID** | | | |
| Agree | 131 (33.2) | 107 (32.7) | 141 (38.6) |
| Disagree | 239 (60.7) | 200 (61.2) | 195 (53.4) |
| Don't know | 24 (6.1) | 20 (6.1) | 29 (8.0) |
| **I have had difficulties concentrating because of the coronavirus/COVID** | | | |
| Agree | 150 (38.1) | 132 (40.4) | 148 (40.5) |
| Disagree | 215 (54.6) | 179 (54.7) | 190 (52.1) |
| Don't know | 29 (7.4) | 16 (4.9) | 27 (7.4) |
| **Thinking about the coronavirus/COVID makes me anxious** | | | |
| Agree | 224 (56.8) | 191 (58.4) | 205 (56.2) |
| Disagree | 146 (37.1) | 120 (36.7) | 134 (36.7) |
| Don't know | 24 (6.1) | 16 (4.9) | 26 (7.1) |
| **I am feeling overwhelmed by the coronavirus/COVID** | | | |
| Agree | 174 (44.2) | 126 (38.5) | 159 (43.6) |
| Disagree | 177 (44.9) | 169 (51.7) | 170 (46.6) |
| Don't know | 43 (10.9) | 32 (9.8) | 36 (9.9) |
| **I am worried about money because of the coronavirus/COVID** | | | |
| Agree | 246 (62.4) | 224 (68.5) | 256 (70.1) |
| Disagree | 126 (32.0) | 93 (28.4) | 94 (25.7) |
| Don't know | 22 (5.6) | 10 (3.1) | 15 (4.1) |

[1] COVID-19 related distress adapted from [25].

**STROBE Statement—checklist of items that should be included in reports of observational studies**

| | Item No | Recommendation | Page number |
|---|---|---|---|
| **Title and abstract** | 1 | (*a*) Indicate the study's design with a commonly used term in the title or the abstract | Abstract |
| | | (*b*) Provide in the abstract an informative and balanced summary of what was done and what was found | Abstract |
| **Introduction** | | | |
| Background/rationale | 2 | Explain the scientific background and rationale for the investigation being reported | Background Pages 4-5 |
| Objectives | 3 | State specific objectives, including any prespecified hypotheses | Background Page 5 paragraph 4 |
| **Methods** | | | |
| Study design | 4 | Present key elements of study design early in the paper | Methods Page 6 "Study Design and Participants" |
| Setting | 5 | Describe the setting, locations, and relevant dates, including periods of recruitment, exposure, follow-up, and data collection | Methods Pages 6-7 |
| Participants | 6 | (*a*) *Cohort study*—Give the eligibility criteria, and the sources and methods of selection of participants. Describe methods of follow-up<br>*Case-control study*—Give the eligibility criteria, and the sources and methods of case ascertainment and control selection. Give the rationale for the choice of cases and controls<br>*Cross-sectional study*—Give the eligibility criteria, and the sources and methods of selection of participants | Methods Pages 6-7 |
| | | (*b*) *Cohort study*—For matched studies, give matching criteria and number of exposed and unexposed<br>*Case-control study*—For matched studies, give matching criteria and the number of controls per case | NA |
| Variables | 7 | Clearly define all outcomes, exposures, predictors, potential confounders, and effect modifiers. Give diagnostic criteria, if applicable | Methods Pages 8-12 |
| Data sources/ measurement | 8 | For each variable of interest, give sources of data and details of methods of assessment (measurement). Describe comparability of assessment methods if there is more than one group | Methods Pages 7-10 |
| Bias | 9 | Describe any efforts to address potential sources of bias | Methods Pages 10-12 |
| Study size | 10 | Explain how the study size was arrived at | Methods Page 6 |
| Quantitative variables | 11 | Explain how quantitative variables were handled in the analyses. If applicable, describe which groupings were chosen and why | Methods Pages 7-12 |
| Statistical methods | 12 | (*a*) Describe all statistical methods, including those used to control for confounding | Methods Pages 10-12 |
| | | (*b*) Describe any methods used to examine subgroups and interactions | NA |
| | | (*c*) Explain how missing data were addressed | Methods Page 8, 10 |
| | | (*d*) *Cohort study*—If applicable, explain how loss to follow-up was addressed | Methods Page 7-8 |
| | | (*e*) Describe any sensitivity analyses | NA |
| **Results** | | | |

| | | | |
|---|---|---|---|
| Participants | 13 | (a) Report numbers of individuals at each stage of study—eg numbers potentially eligible, examined for eligibility, confirmed eligible, included in the study, completing follow-up, and analysed | Figure 1, and throughout tables |
| | | (b) Give reasons for non-participation at each stage | NA |
| | | (c) Consider use of a flow diagram | Figure 1 |
| Descriptive data | 14 | (a) Give characteristics of study participants (eg demographic, clinical, social) and information on exposures and potential confounders | p. 13 and Table 1 |
| | | (b) Indicate number of participants with missing data for each variable of interest | Tables 1 and 2 |
| | | (c) *Cohort study*—Summarise follow-up time (e.g., average and total amount) | Figure 1 and Table 1 |
| Outcome data | 15 | *Cohort study*—Report numbers of outcome events or summary measures over time | Table 1 |
| | | *Case-control study*—Report numbers in each exposure category, or summary measures of exposure | NA |
| | | *Cross-sectional study*—Report numbers of outcome events or summary measures | NA |
| Main results | 16 | (*a*) Give unadjusted estimates and, if applicable, confounder-adjusted estimates and their precision (eg, 95% confidence interval). Make clear which confounders were adjusted for and why they were included | Tables 3, 4, 5 summarized in pp. 13-14 |
| | | (*b*) Report category boundaries when continuous variables were categorized | NA |
| | | (*c*) If relevant, consider translating estimates of relative risk into absolute risk for a meaningful time period | NA |
| Other analyses | 17 | Report other analyses done—eg analyses of subgroups and interactions, and sensitivity analyses | NA |
| **Discussion** | | | |
| Key results | 18 | Summarise key results with reference to study objectives | pp. 15-16 |
| Limitations | 19 | Discuss limitations of the study, taking into account sources of potential bias or imprecision. Discuss both direction and magnitude of any potential bias | pp.19 |
| Interpretation | 20 | Give a cautious overall interpretation of results considering objectives, limitations, multiplicity of analyses, results from similar studies, and other relevant evidence | pp. 15-19 |
| Generalisability | 21 | Discuss the generalisability (external validity) of the study results | pp.19 |
| **Other information** | | | |
| Funding | 22 | Give the source of funding and the role of the funders for the present study and, if applicable, for the original study on which the present article is based | Title page |
